# Prevalence of COVID-19 and long COVID - Results from the 2022 Behavioral Risk Factor Surveillance System, 50 states

**DOI:** 10.1101/2023.09.29.23296352

**Authors:** Mary L. Adams, Joseph Grandpre

**Affiliations:** On Target Health Data LLC, Suffield, Connecticut, USA; Wyoming Department of Health, Cheyenne, Wyoming, USA

## Abstract

Recent MMWR results estimate long COVID prevalence at 6.0% in June 2023, while the percentage of those with COVID reporting long COVID was 11.0%. The 2022 Behavioral Risk Factor Surveillance System addressed COVID (positive test) and long COVID (symptoms lasting ≥3 months) in a population-based sample from each state and DC. Results for 385,617 adults indicated 34.4% had ever had COVID, 21.9% of whom reported long COVID, representing 7.4% of all adults. State rates ranged from 25.4% - 40.8% for COVID and 4.1%- 11.1% for long COVID. Groups with high rates for both included women, younger adults, those with children in the household, plus those reporting obesity, asthma, chronic obstructive pulmonary disease (COPD), diabetes, or cardiovascular disease (CVD). Highest adjusted odds ratios for COVID were 2.34 (95% CI 2.20-2.49) for age 18-24 years vs. age 65+ while for long COVID it was 2.81 (2.53-3.13) for 3+ of the 5 conditions. Most frequently reported problems for those with long COVID were fatigue (26.0%), shortness of breath (18.8%), loss of taste or smell (17.2%), and memory problems (9.9%). Results show the need for state-based data and suggest a focus on younger adults is needed to address COVID and long COVID.

## Introduction

Recent results estimate long COVID prevalence dropped from 7.5% in June 2022 to 6.0% in June 2023, while the percentage of those with COVID reporting long COVID dropped from 18.9% to 11.0% (*1*). The Behavioral Risk Factor Surveillance System (BRFSS) is an ongoing state-based telephone survey of randomly selected non-institutionalized adults that collects data on a range of health topics (*2*). This study used data from the 2022 BRFSS which included questions on COVID and long COVID to provide state-based rates of both along with information on groups at higher risk.

## Methods

BRFSS data are publicly available from all 50 states and DC (*3*). The COVID questions were 1) “Has a doctor, nurse, or other health professional ever told you that you tested positive for COVID 19?” with possible responses of yes, no, or “Tested positive using home test without health professional” and 2) “Did you have any symptoms lasting 3 months or longer that you did not have prior to having coronavirus or COVID-19?” This was considered long COVID. Other measures included age, race, gender, income, children in the household, census region (Northeast, Midwest, South and West), smoking, e-cigarette use, and any HIV risk factor (recent injected drug use, STD treatment or exchanging sex for money or drugs). A composite measure of 5 chronic conditions (obesity, asthma, cardiovascular disease (CVD), diabetes, and COPD (Chronic Obstructive Pulmonary Disease) found to be associated with US hospitalizations for COVID (*4, 5*) was also included. Data were weighted to adjust for the probability of selection and to reflect the adult population of each state by age group, race/ethnicity, education level, marital status, and home ownership. Stata version 18.0 (StataCorp LLC, College Station TX) was used to account for the complex sample design of the BRFSS in unadjusted analysis and also controlled for the listed factors in logistic regression. Missing values for any measure were excluded from analysis. The median response rate for the 50 states plus DC for land line and cell phone surveys combined was 45.1% ranging from 36.2% in CA to 66.8% in SD. A total of 385,617 respondents were included in the analysis with state N’s ranging from 3,188 in NV to 26,152 in WA and a median N of 7, 473.

## Results

Results (Table 1) indicate that 34.4% of respondents had ever had a positive test for COVID, ranging from 25.4% in OR to 40.8% in MS. One in five adults with COVID (21.9%) reported long COVID, ranging from 14.2% in HI to 29.2% in WV, representing 7.4% of all adults, with a range from 4.1% in HI to 11.1% in WV. Higher rates for COVID were found for women with children in the household (42.8%), current e-cigarette users (41.5%), (but not for smokers), younger adults (39.4-40.7% for 18-44-year-old age groups), income >$75K (≥39.5%), extreme obesity (39.8%), HIV risk (43.2%), and current asthma (40.0%). Higher rates for long COVID were reported by most of these same groups plus the South and Midwest regions (7.8% and 7.7% respectively), adults on Medicaid (9.4%), and those with more of the 5 chronic conditions.

**Table 1.**
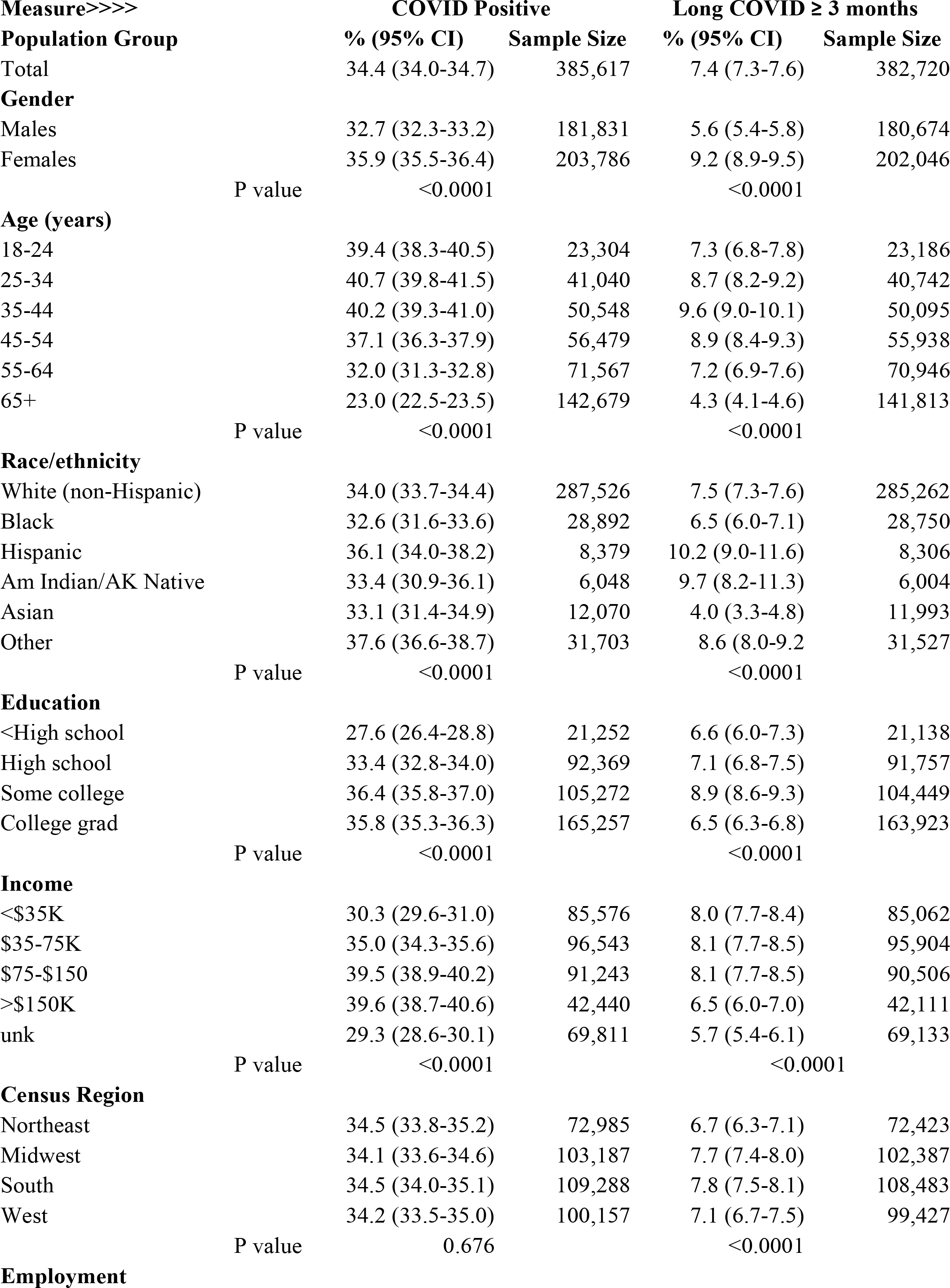
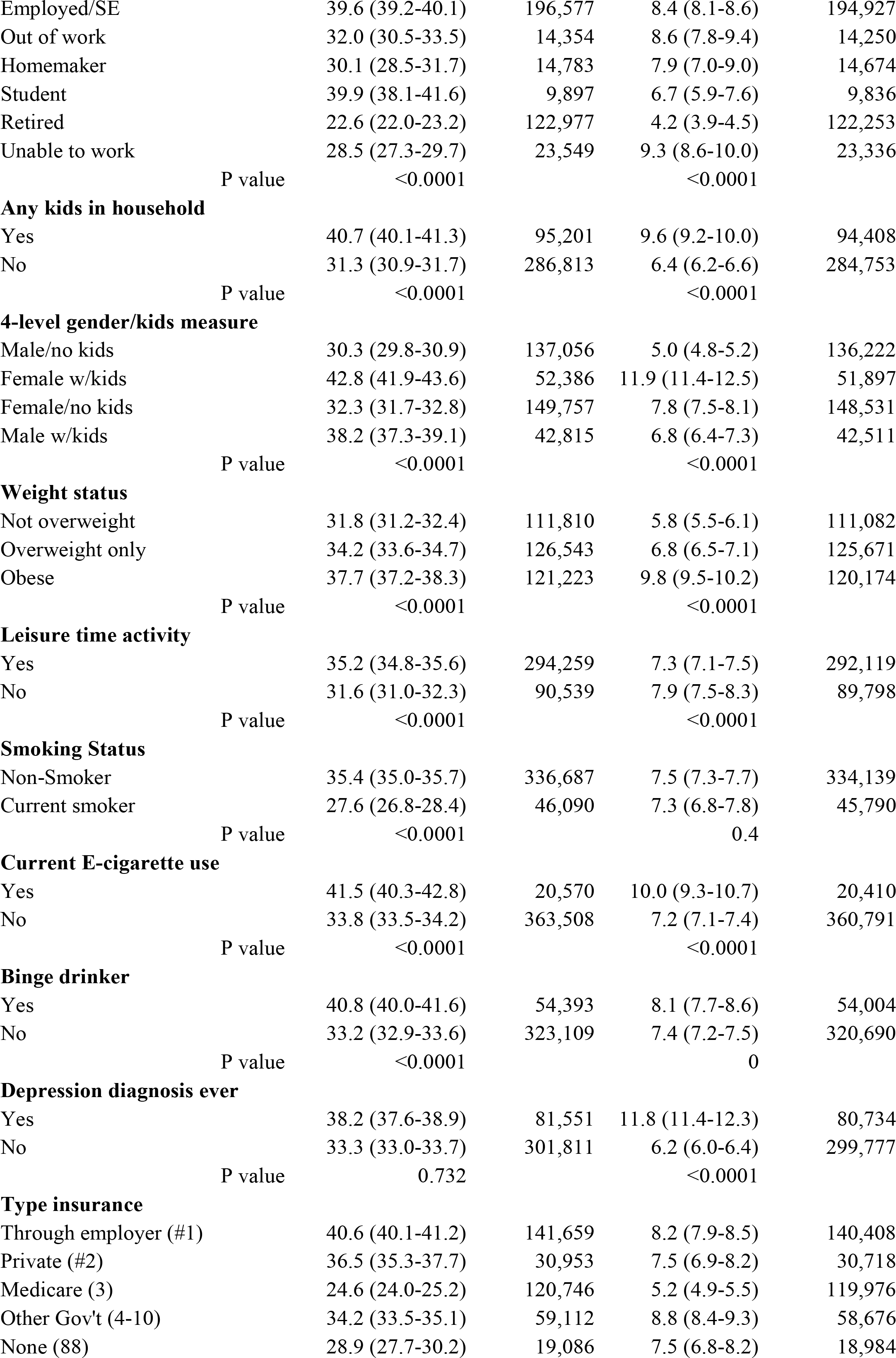
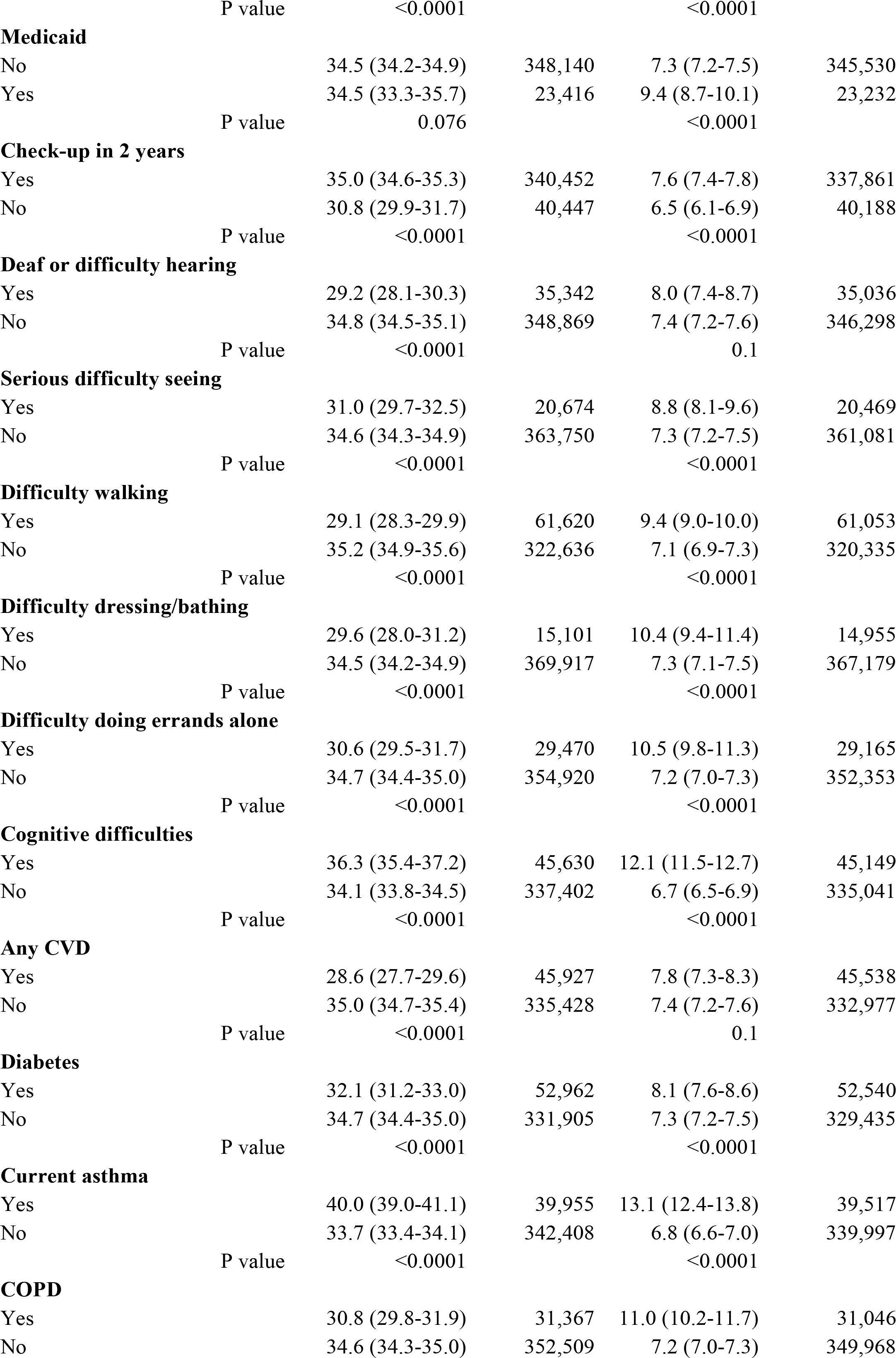
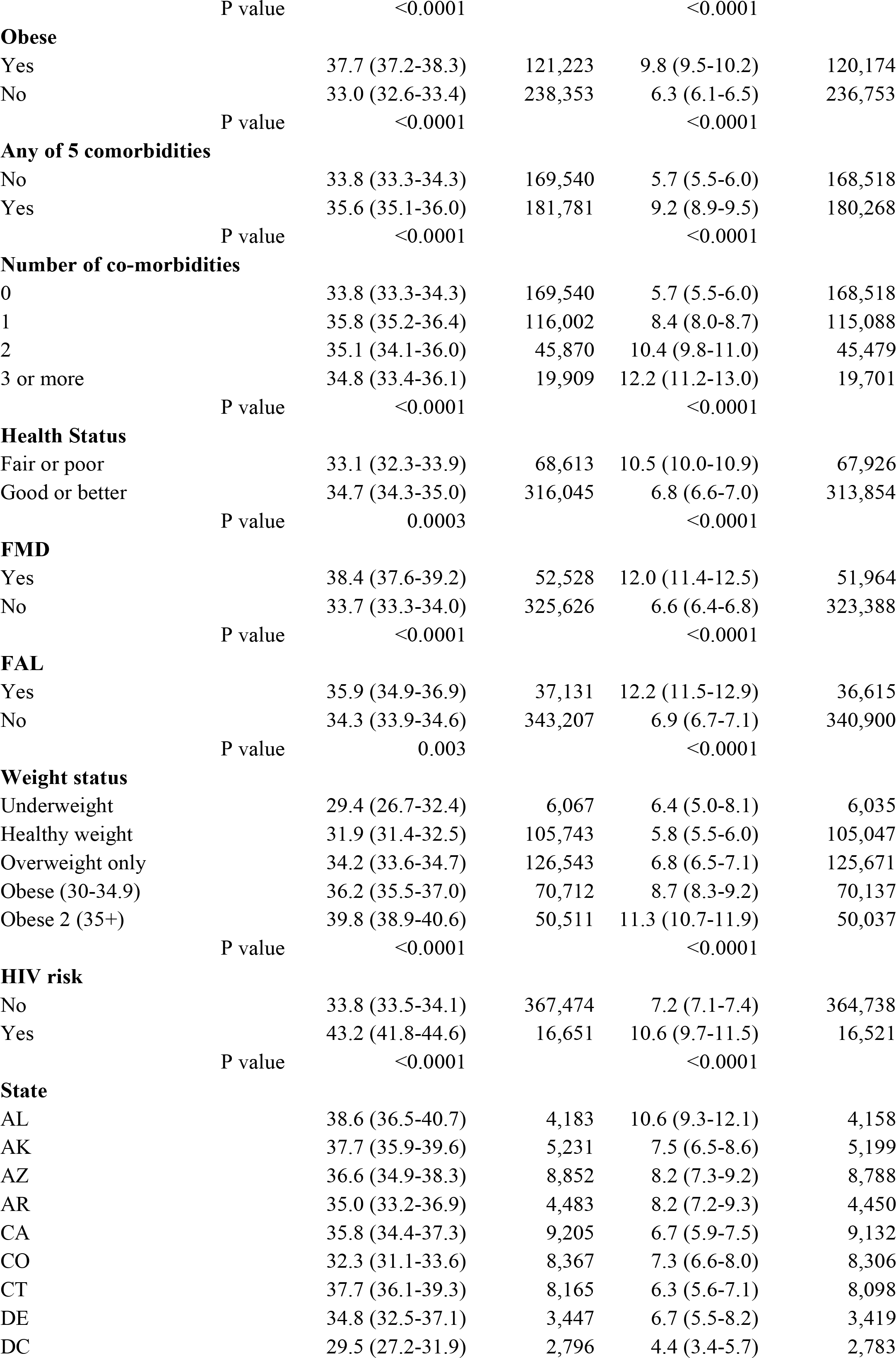
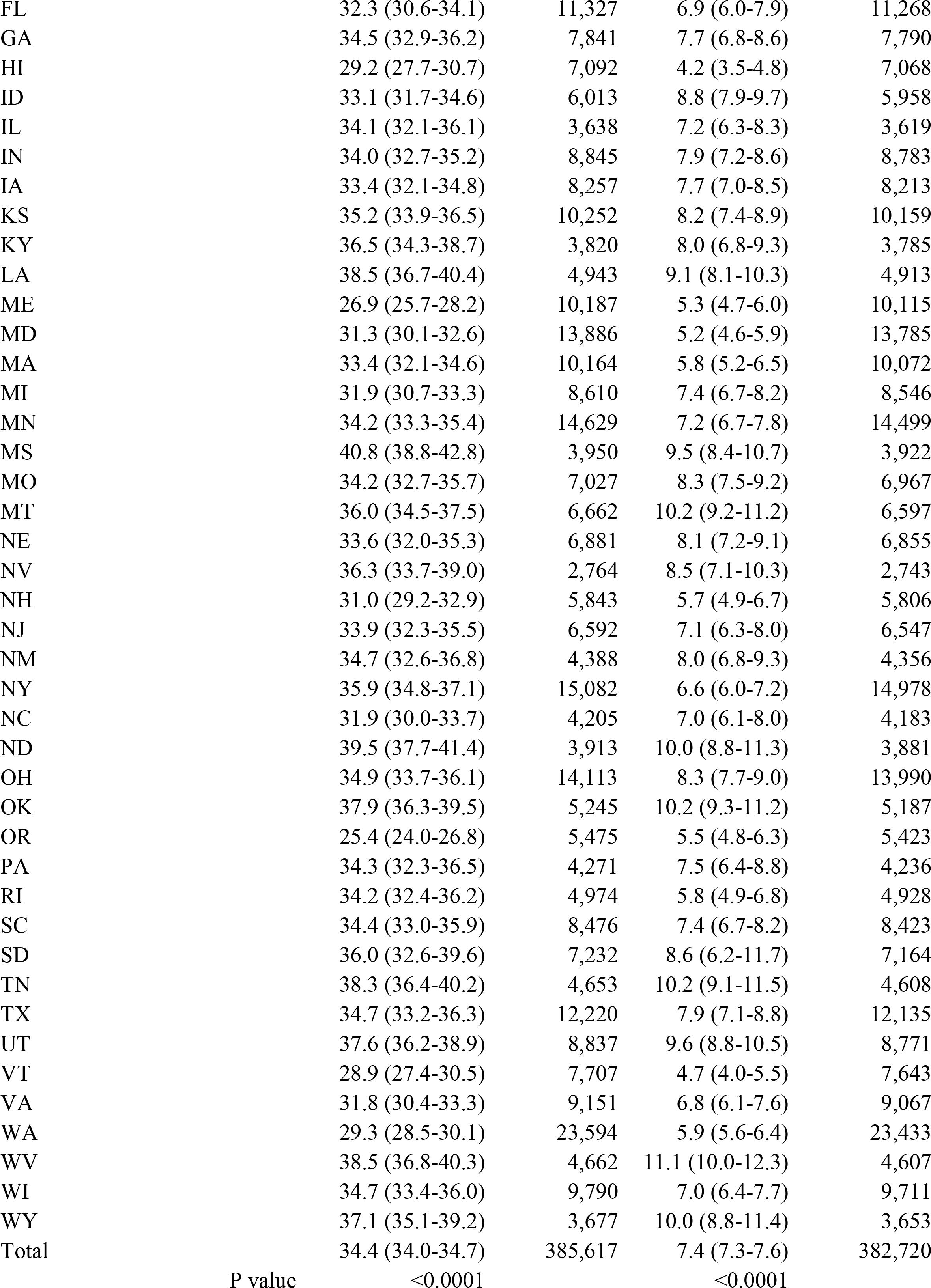
COVID and Long COVID, 2022 Behavioral Risk Factor Surveillance System, 50 states. N=338,465 for COVID, 336,082 for long COVID. Results from weighted analysis in Stata, State N’s ranged from 3,188 in NV-26,152 in WA, median=7,473. Comorbidities: obesity, diabetes, cardiovascular disease (CVD), asthma, or chronic obstructive pulmonary disease (COPD).

Among adults with none of the 5 comorbidities, 5.7% reported long COVID compared with 12.1% of those with 3 or more. Results were confirmed by logistic regression using models including these measures plus demographics (Table 2). Other factors with higher rates among those reporting long COVID such as cognitive difficulties (12.1%), depression (11.8%), or a cost barrier to health care (13.0%) might have resulted from long COVID and were not included in logistic regression models. The problems most frequently reported by those with long COVID were fatigue (26.0%), breathing problems (18.8%), loss of taste or smell (17.2%), and memory problems (9.9%), while 22.9% reported some other problem and 5.1% reported no long-term symptoms that limited activities.

**Table 2.** Results of logistic regression, controlled for the measures listed, 2022 Behavioral Risk Factor Surveillance System, 50 states & DC, N=338,465 for COVID, 336,082 for long COVID. State N’s ranged from 3,188 in NV-26,152 in WA, median=7,473. HI with lowest rate for COVID was chosen as referent for both measures. COVID risks=obesity, diabetes, cardiovascular disease, chronic obstructive pulmonary disease, asthma.

| Outcome>>> | COVID Positive |  | Long COVID ≥3 months |  |
| --- | --- | --- | --- | --- |
| Group | AOR (95% CI) | P value | AOR (95% CI) | P value |
| Females w/kids v M, no kids | 1.45 (1.38-1.52) | <0.0001 | 2.22 (2.04-2.41) | <0.0001 |
| F no kids v M no kids | 1.20 (1.15-1.24) | <0.0001 | 1.76 (1.65-1.89) | <0.0001 |
| M w/kids v M no kids | 1.15 (1.09-1.20) | <0.0001 | 1.18 (1.08-1.30) | <0.0001 |
| 55-64 years v 65+ | 1.49 (1.42-1.57) | <0.0001 | 1.79 (1.64-1.96) | <0.0001 |
| 45-54 years v 65+ | 1.79 (1.70-1.89) | <0.0001 | 2.24 (2.04-2.47) | <0.0001 |
| 35-44 years v 65+ | 2.00 (1.89-2.12) | <0.0001 | 2.41 (2.17-2.68) | <0.0001 |
| 25-34 years v 65+ | 2.21 (2.10-2.34) | <0.0001 | 2.35 (2.13-2.61) | <0.0001 |
| 18-24 years v 65+ | 2.34 (2.20-2.49) | <0.0001 | 2.21 (1.97-2.48) | <0.0001 |
| Black v non-Hispanic white | 0.87 (0.83-0.92) | <0.0001 | 0.73 (0.67-0.81) | <0.0001 |
| Hispanic v non-Hispanic white | 0.98 (0.89-1.08) | 0.655 | 1.23 (1.05-1.43) | 0.009 |
| Am. Indian v non_Hispanic white | 0.94 (0.83-1.07) | 0.358 | 1.13 (0.92-1.39) | 0.241 |
| Asian v. non_Hispanic white | 0.85 (0.78-0.93) | 0.001 | 0.53 (0.43-0.66) | <0.0001 |
| Other v. non_Hispanic white | 1.10 (1.05-1.17) | <0.0001 | 1.06 (0.97-1.16) | 0.203 |
| \$25-<\$50K v < \$25K | 1.30 (1.22-1.38) | <0.0001 | 1.12 (1.01-1.24) | 0.03 |
| \$50-<\$75K v < \$25K | 1.51 (1.42-1.61) | <0.0001 | 1.21 (1.09-1.35) | <0.0001 |
| \$75K-<\$100K v < \$25K | 1.66 (1.56-1.78) | <0.0001 | 1.27 (1.13-1.42) | <0.0001 |
| \$100K+ v <\$25K | 1.79 (1.69-1.90) | <0.0001 | 1.08 (0.97-1.20) | 0.159 |
| Unk inc v <\$25K | 1.14 (1.07-1.22) | <0.0001 | 0.88 (0.78-0.98) | 0.022 |
| 1 COVID risk v 0 | 1.18 (1.13-1.22) | <0.0001 | 1.51 (1.42-1.62) | <0.0001 |
| 2 COVID risk v 0 | 1.37 (1.30-1.44) | <0.0001 | 2.21 (2.04-2.39) | <0.0001 |
| 3 COVID risk v 0 | 1.56 (1.45-1.67) | <0.0001 | 2.81 (2.53-3.13) | <0.0001 |
| HIV risk* v no | 1.22 (1.15-1.30) | <0.0001 | 1.29 (1.17-1.44) | <0.0001 |
| E-cigarettes v no | 1.12 (1.06-1.19) | <0.0001 | 1.18 (1.08-1.30) | <0.0001 |
| State |  |  |  |  |
| AL v HI | 1.46 (1.29-1.67) | <0.0001 | 2.27 (1.77-2.91) | <0.0001 |
| AK v HI | 1.25 (1.11-1.41) | <0.0001 | 1.38 (1.07-1.76) | 0.012 |
| AZ v HI | 1.29 (1.15-1.46) | <0.0001 | 1.54 (1.21-1.95) | <0.0001 |
| AR v HI | 1.24 (1.09-1.40) | 0.001 | 1.55 (1.21-1.97) | <0.0001 |
| CA v HI | 1.27 (1.14-1.42) | <0.0001 | 1.45 (1.15-1.83) | 0.002 |
| CO v HI | 1.00 (0.90-1.11) | 0.942 | 1.47 (1.18-1.83) | 0.001 |
| CT v HI | 1.36 (1.21-1.53) | <0.0001 | 1.32 (1.05-1.67) | 0.02 |
| DE v HI | 1.21 (1.05-1.39) | 0.009 | 1.39 (1.04-1.86) | 0.028 |
| DC v HI | 0.89 (0.76-1.03) | 0.108 | 0.96 (0.69-1.34) | 0.814 |
| FL v HI | 1.12 (0.99-1.26) | 0.084 | 1.45 (1.13-1.86) | 0.003 |
| GA v HI | 1.22 (1.09-1.38) | 0.001 | 1.62 (1.28-2.05) | <0.0001 |
| ID v HI | 1.10 (0.98-1.23) | 0.105 | 1.75 (1.40-2.20) | <0.0001 |
| IL v HI | 1.15 (1.01-1.31) | 0.036 | 1.49 (1.16-1.92) | 0.002 |
| IN v HI | 1.15 (1.03-1.28) | 0.011 | 1.54 (1.24-1.91) | <0.0001 |
| IA v HI | 1.10 (0.98-1.22) | 0.099 | 1.47 (1.17-1.83) | 0.001 |
| KS v HI | 1.16 (1.04-1.29) | 0.006 | 1.60 (1.28-1.99) | <0.0001 |
| KY v HI | 1.27 (1.11-1.46) | 0.001 | 1.47 (1.13-1.91) | 0.005 |
| LA v HI | 1.42 (1.26-1.60) | <0.0001 | 1.73 (1.36-2.20) | <0.0001 |
| ME v HI | 0.87 (0.77-0.97) | 0.011 | 1.05 (0.82-1.33) | 0.711 |
| MD v HI | 1.01 (0.91-1.12) | 0.854 | 1.04 (0.82-1.31) | 0.748 |
| MA v HI | 1.14 (1.02-1.26) | 0.019 | 1.21 (0.96-1.51) | 0.101 |
| MI v HI | 1.07 (0.96-1.19) | 0.251 | 1.47 (1.17-1.84) | 0.001 |
| MN v HI | 1.14 (1.03-1.26) | 0.011 | 1.14 (1.16-1.78) | 0.001 |
| MS v HI | 1.65 (1.46-1.88) | <0.0001 | 1.98 (1.55-2.52) | <0.0001 |
| MO v HI | 1.18 (1.05-1.32) | 0.005 | 1.68 (1.34-2.10) | <0.0001 |
| MT v HI | 1.27 (1.13-1.42) | <0.0001 | 2.03 (1.62-2.54) | <0.0001 |
| NE v HI | 1.10 (0.98-1.24) | 0.107 | 1.56 (1.24-1.96) | <0.0001 |
| NV v HI | 1.24 (1.06-1.44) | 0.006 | 1.75 (1.31-2.33) | <0.0001 |
| NH v HI | 0.99 (0.87-1.12) | 0.861 | 1.14 (0.87-1.49) | 0.335 |
| NJ v HI | 1.16 (1.03-1.30) | 0.011 | 1.63 (1.28-2.07) | <0.0001 |
| NM v HI | 1.17 (1.02-1.33) | 0.026 | 1.43 (1.09-1.86) | 0.009 |
| NY v HI | 1.29 (1.17-1.43) | <0.0001 | 1.43 (1.15-1.78) | 0.002 |
| NC v HI | 1.13 (1.00-1.28) | 0.058 | 1.53 (1.19-1.95) | 0.001 |
| ND v HI | 1.42 (1.26-1.61) | <0.0001 | 1.99 (1.57-2.52) | <0.0001 |
| OH v HI | 1.19 (1.08-1.33) | 0.001 | 1.57 (1.27-1.94) | <0.0001 |
| OK v HI | 1.34 (1.20-1.51) | <0.0001 | 1.93 (1.55-2.41) | <0.0001 |
| OR v HI | 0.76 (0.68-0.86) | <0.0001 | 1.05 (0.82-1.35) | 0.676 |
| PA v HI | 1.19 (1.03-1.36) | 0.014 | 1.47 (1.12-1.93) | 0.005 |
| RI v HI | 1.14 (1.00-1.29) | 0.043 | 1.11 (0.85-1.45) | 0.456 |
| SC v HI | 1.25 (1.12-1.40) | <0.0001 | 1.61 (1.29-2.02) | <0.0001 |
| SD v HI | 1.23 (1.03-1.48) | 0.023 | 1.42 (1.02-1.96) | 0.036 |
| TN v HI | 1.37 (1.21-1.55) | <0.0001 | 2.00 (1.58-2.54) | <0.0001 |
| TX v HI | 1.11 (0.99-1.25) | 0.071 | 1.54 (1.23-1.92) | <0.0001 |
| UT v HI | 1.20 (1.08-1.34) | 0.001 | 1.82 (1.46-2.25) | <0.0001 |
| VT v HI | 0.95 (0.85-1.07) | 0.413 | 0.96 (0.74-1.25) | 0.753 |
| VA v HI | 1.03 (0.92-1.15) | 0.625 | 1.39 (1.10-1.75) | 0.005 |
| WA v HI | 0.90 (0.82-0.98) | 0.021 | 1.15 (0.94-1.41) | 0.164 |
| WV v HI | 1.46 (1.30-1.65) | <0.0001 | 2.14 (1.70-2.69) | <0.0001 |
| WI v HI | 1.18 (1.06-1.32) | 0.002 | 1.33 (1.07-1.66) | 0.011 |
| WY v HI | 1.32 (1.16-1.50) | <0.0001 | 2.05 (1.61-2.62) | <0.0001 |
| COVID risks if entered separately |  |  |  |  |
| Obesity | 1.21 (1.17-1.25) | <0.0001 | 1.42 (1.34-1.51) | <0.0001 |
| Diabetes | 1.14 (1.08-1.19) | <0.0001 | 1.19 (1.09-1.29) | <0.0001 |
| Cardiovascular disease | 1.06 (1.01-1.13) | 0.03 | 1.28 (1.17-1.39) | <0.0001 |
| COPD | 1.01 (0.95-1.08) | 0.76 | 1.46 (1.31-1.62) | <0.0001 |
| Asthma | 1.25 (1.19-1.32) | <0.0001 | 1.65 (1.52-1.78) | <0.0001 |

Abbreviations: AOR: adjusted odds ratio; COPD: chronic obstructive pulmonary disease;
Unk: unknown.
\* HIV risk: Any in past year: Injected a non-prescribed drug; treated for a sexually transmitted disease; exchanged money or drugs for sex.

## Discussion

Population based data from the BRFSS provide new information on COVID and long COVID. First is the wide range in rates of both COVID and long COVID among the states which translated to regional differences only for long COVID and remained when results were controlled for all the measures included in logistic regression. Five states had adjusted odds ratios of 2.0 or higher for long COVID. Second, one in five adults with a positive COVID test reported long COVID, again showing a wide range across states. Third, some groups at risk were different from those identified earlier. High risk groups for COVID and long COVID appear to reflect younger age groups with higher income compared with groups identified early in the pandemic based on hospitalizations (*4, 5*). The elderly/retired, smokers, low-income adults, and most minorities appear at low risk. However, those reporting any of obesity, diabetes, COPD, CVD, and asthma – conditions identified as increasing risk of COVID hospitalizations (*4*) – generally reported higher rates of both COVID and long COVID although results for the separate measures differed. The role of obesity in COVID risk appears to be present even when the outcome is cases rather than deaths (*6*). The prevalence rate for long COVID of 7.4% among all adults is consistent with that recently reported (*1*), but much higher than the 1.7% in another study where long COVID was defined as symptoms lasting 2 months (*7*) and not as “ever’.

However, these current results for the most common problems reported for long COVID are similar to those from a meta-analysis (*8*). Another factor to consider in risk is the number of respondents reporting the measure. For example, obesity and reporting a risk for HIV appear to increase rates of COVID and long COVID approximately the same amount (Tables 1 & 2) but there were about seven times as many respondents with obesity as reporting HIV risk (121,000 vs. 17,000).

Limitations: There are at least four limitations to this study. First, because the BRFSS only surveys households, among the institutions the survey omits are nursing homes and prisons which appeared to have high rates of COVID especially early in the pandemic. Thus, results may underestimate the true rates of COVID and long COVID. Second, results are self-reported and except as noted for COVID are not based on an actual test or diagnosis; the implications of this limitation are unknown. Third, the lack of a measure of hypertension on the survey for 2022 meant that the composite measure of comorbidities lacked a key component (*5*). Fourth, survey results can’t distinguish cause and effect so results indicating higher rates of poor health, cognitive disability, frequent activity limitation, and reporting a cost barrier to health care for those reporting long COVID only indicate an association so were omitted from logistic regression.

Implications for public health: These results show that younger adults and especially women with children in the household, appear to be a high-risk group for COVID and long COVID. Results also confirm that 5 of the 6 comorbidities that were found to increase risk of hospitalization for COVID are also associated with long COVID which affected one-fifth of adults with COVID. A potential key factor not included in this study is vaccines (*9*) where data are only available for about half the states. The wide range in rates of COVID and long COVID across states even when controlled for a range of demographic and health measures should be recognized and addressed, along with regional differences in long COVID. The finding of long-term memory problems reported by 9.9% of those with long COVID along with higher rates of both COVID and long COVID reported by respondents reporting cognitive difficulties, underscores the need for continued monitoring of cognitive health among all ages and not limited to ages 45+.

## Data Availability

Data used are available at: https://www.cdc.gov/brfss/annual_data/annual_2022.html. Data produced in study that are not included in paper are available upon reasonable request to authors.

